# Disability patterns in multiple sclerosis: a meta-analysis on PIRA and RAW in the real world context

**DOI:** 10.1101/2024.04.08.24305472

**Authors:** Luca Prosperini, Serena Ruggieri, Shalom Haggiag, Carla Tortorella, Claudio Gasperini

## Abstract

**Background:** The confirmed disability accrual (CDA) due to multiple sclerosis (MS) is driven by two factors: relapse-associated worsening (RAW) and progression independent of relapse activity (PIRA). However, accurate estimations of these phenomena in the real-world setting are lacking. This study aims at summarizing current evidence on RAW and PIRA, including associated factors, through a quantitative synthesis of real-world studies.

**Methods:** Scientific databases were searched to identify real-world studies published until December 31, 2023, reporting how many patients experienced RAW and PIRA (events of interest). Random-effects meta-analyses, subgroup analyses and meta-regression models were ran to provide pooled estimates of RAW and PIRA events, and to identify their potential moderators (PROSPERO registration: CRD42024503895).

**Results:** Eighteen articles met the eligibility criteria, with a pooled sample size of 52,667 patients followed for 2.4 to 12.1 years (415,825 patient-years). Pooled event rates for RAW and PIRA were 1.6 and 3.1 per 100 patient-years, respectively. Less RAW events were found in patient cohorts under high-efficacy disease-modifying treatments (β=–0.031, p=0.007), while PIRA events were directly related to older age (β=0.397, p=0.027), predicting ≥6 PIRA events per 100 patient-years at an age ≥54 years. Additionally, we found significant differences in PIRA event rates according to the criteria adopted to define CDA.

**Discussion:** PIRA accounts for most CDA events in the real-world setting, even at the earlier disease stages, whereas RAW represents a less frequent phenomenon, likely due to effective treatments. However, the detection and statistical analysis of PIRA outcomes pose challenges, raising the risk of biased interpretation.

**What is already known on this topic:** Irreversible accumulation of disability in multiple sclerosis stems from two distinct yet not mutually exclusive phenomena: relapse-associated worsening (RAW) and progression independent of relapse activity (PIRA).

**What this study adds:** In this meta-analysis including data of 52,667 patients followed for 2.4 to 12.1 years, the pooled event rates were 1.6 and 3.1 per 100 patient-years for RAW and PIRA, respectively. High-efficacy treatment strongly suppresses RAW but not PIRA, which is instead associated with age and definition criteria.

**How this study might affect research, practice or policy:** Although PIRA accounts for most disability events in MS, its detection in real-world setting is necessarily inaccurate and its statistical analysis is challenging.

## INTRODUCTION

Irreversible accumulation of disability in multiple sclerosis (MS) stems from two distinct yet not mutually exclusive phenomena: relapse-associated worsening (RAW) and progression independent of relapse activity (PIRA). RAW involves a step-wise increase in impairment due to incomplete recovery from a relapse, while PIRA entails a relentless accumulation of permanent disability unrelated to relapses [1]. Historically, relapses were considered the main source of permanent disability in early disease phases, when focal inflammation dominated over diffuse inflammation and neurodegeneration. Disease-modifying treatments (DMTs) exert their strongest effects in these early stages, and post-relapse accumulation of disability is attributed to compensative mechanism failure or lesion location in critical regions of the central nervous system (CNS) [2]. In later phases, characterized as ‘smouldering MS’ [3], predominant diffuse inflammation and neurodegeneration lead to insidious disability progression, while DMTs becoming largely ineffective. With disease progression and aging-related processes, the CNS loses compensatory abilities, resulting in brain network collapse [4]. A ‘two-stage disease’ hypothesis proposes two clearly distinct phases, earlier inflammatory and later neurodegenerative, corresponding to relapses and disability progression, respectively [5].

Contrary to this perspective, recent data from post-hoc analysis of randomized clinical trials (RCTs) and observational studies show that RAW and PIRA can co-occur in the same individuals [1],[6], and that PIRA events may occur soon after the first demyelinating event or in paediatric-onset MS [1],[7]. A recently published systematic review indicated no uniform definition of PIRA in the literature, estimating roughly 5% of yearly PIRA events, contributing to at least 50% of all confirmed disability accrual (CDA) events in typical relapsing-remitting (RR) MS [7]. Another systematic review found variable proportions of patients experiencing PIRA (4 to 24%) based on different definitions and follow-up durations [8].

In this context, we conducted a quantitative synthesis of the currently available real-world evidence on RAW and PIRA, based on CDA as assessed by change in the Expanded Disability Status Scale (EDSS).

Exclusively relying on real-world studies, we avoided composite measures involving clinical measurements that are difficult to implement in daily clinical setting, e.g. timed 25-foot walk test (25-FWT), 9-hole peg test (9-HPT), etc. [6]. Our goal was to provide accurate estimates and explore moderators influencing the RAW and PIRA event rates through subgroup meta-analyses meta-regressions.

## METHODS

### Study Design and Registration

Our systematic review and meta-analysis followed the Preferred Reporting Items for Systematic Reviews and Meta-analyses (PRISMA) statement [9] and is registered in the PROSPERO database (CRD42024503895).

### Search Strategy

To identify studies to include in this meta-analysis, we searched PubMed/Medline and Google Scholar, using combinations of free-text and MeSH terms for articles published until December 31, 2023 as follows: (“relapse associated worsening” OR “RAW”) AND (“progression independent of relapse activity” OR “PIRA” OR “silent progression”) AND (“multiple sclerosis”).

One reviewer ran the search strategy and screened the initial titles after removing duplicates. Two authors independently examined each potential relevant article, using the following criteria as defined by the PI(E)COS framework [10]: (i) Population: persons with MS; (ii) Intervention: not applicable (iii) Comparison: not applicable (iv): Outcomes: RAW and PIRA based on EDSS change; (v) setting: real-world setting. We excluded case reports and case series, articles not written in English and those reporting data from experimental setting (e.g. post-hoc analysis from RCTs).

### Data Extraction

Two authors independently extracted data, with discrepancies resolved by a third author. The following information was extracted from each article: first author, publication year, total sample size, study follow-up (years), mean age (years), mean time elapsed from the clinical onset (years), median EDSS score, type of DMTs, number of patients who experienced RAW and PIRA (outcomes of interest).

Given that there is no uniform agreed-upon definition for CDA, especially in the context of PIRA, we also extracted the following criteria from selected articles: baseline score, event score, confirmation score and sustained score [7]. The baseline score refers to the EDSS score at study entry (fixed) or the last confirmed reference score (roving) [11]. The event score indicates the stepwise disability increase reflecting a clinically relevant deterioration, adjusted for the non-linear distribution of EDSS: ≥1.5-step increase for a reference EDSS = 0; ≥1.0-step increase for a reference EDSS of 1.0 to either 5.0 or 5.5; 0.5-step increase for a reference EDSS of either ≥5.5 or 6.0. The confirmation score is the next assessment at a pre-specified period after the event, usually taking place at 3, 6 or 12 months after the initial disability increase. The sustained score is equivalent to the last on-study follow-up, sensibly at least 12 or even 24 months apart from start of CDA. When reported data were insufficient for the analysis, we proactively contacted the lead author of the study to request access to additional information (see Acknowledgements).

Quality assessment of individual studies and across studies was performed by the Newcastle–Ottawa Scale (NOS) [12].

### Statistical analyses

We computed the RAW and PIRA event rates per patient-years, calculated as the sample size multiplied by the study follow-up duration expressed in years. The pooled effect size (ES), with its relative 95% confidence intervals (CIs), was estimated by random-effects models based on the empirical Bayes method. Between-study heterogeneity and inconsistency were assessed by the Cochran’s Q-value and I^2^ index, respectively; we considered an I^2^ ≤25% as marginal, 25–75% as moderate and ≥75% as substantial inconsistency. Risk of publication bias was assessed by the Kendall’s *τ* and *Z* Egger’s tests.

Given the expected intra- and inter-study heterogeneity and in order to identify baseline variables (moderators) potentially influencing both the outcomes of interest, we fitted meta-regression equations and subgroup meta-analyses on arcsine square root-transformed event rates for RAW and PIRA. The arcsine-based ES conversion has many advantages over other transformations, including greater accuracy when the events of interest are rare, variance-stabilizing property, no need to either exclude studies with zero events or add arbitrary increments. As arcsine-transformed estimations are difficult to interpret, we provided graphical charts with effect size estimated as crude event rates, as appropriate.

Regression coefficients (β) with their corresponding 95% confidence intervals (CIs) were also reported for all variables entered in models as moderators; such variables were previously *log*-transformed to met the normality assumption. The coefficient of determination (R^2^) was used to assess the goodness of fit for each weighted model. Two-tailed p-values <0.05 were considered significant.

## RESULTS

### Search findings

Our initial search yielded 124 articles, and after title and abstract screening, we identified 24 articles for eligibility (**<u>FIG. 1</u>**). Of these, 18 articles [13]–[30] published from 2020 to 2023 met the eligibility criteria for this meta-analysis. Seven articles were excluded after full-text reading; four were based on post-hoc analysis of RCTs [1],[6],[31],[32] and three did not report complete data on the events of interest [33]–[35].

**FIG. 1.**
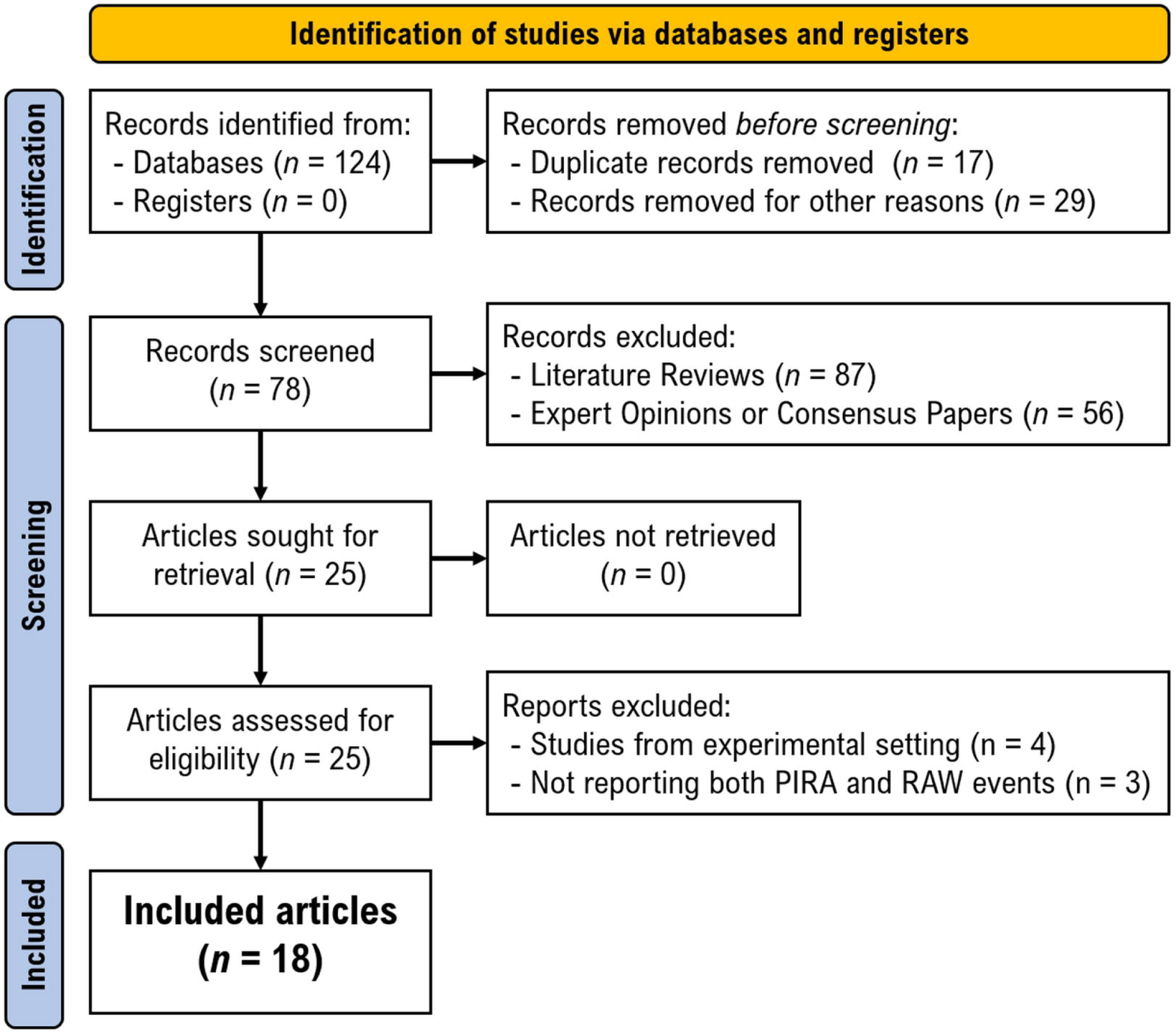
Preferred reporting items for systematic reviews and meta-analyses (PRISMA) flow diagram for study selection. *MS: multiple sclerosis*

The selected studies were conducted in Germany [17],[19],[21],[22],[24], Italy [18],[25]–[27], Switzerland (k = 4) [13],[15],[23],[30], Austria [14], Poland [20], Spain [29], Sweden [28], USA [13], International Registry [16]. Two articles reported composite PIRA based on a combination of EDSS score with either the 25-FWT and 9-HPT [22] or the symbol digit modalities test (SDMT) [14]. Two articles included different patient cohorts that were analysed separately way [13],[23], resulting in k = 20 statistical units.

All selected articles scored 5 or 6 according to the NOS, indicating a good methodological quality; notably, we did not consider the comparability domain in the absence of subgroup comparisons (**<u>TABLE 1</u>**).

**TABLE 1.**
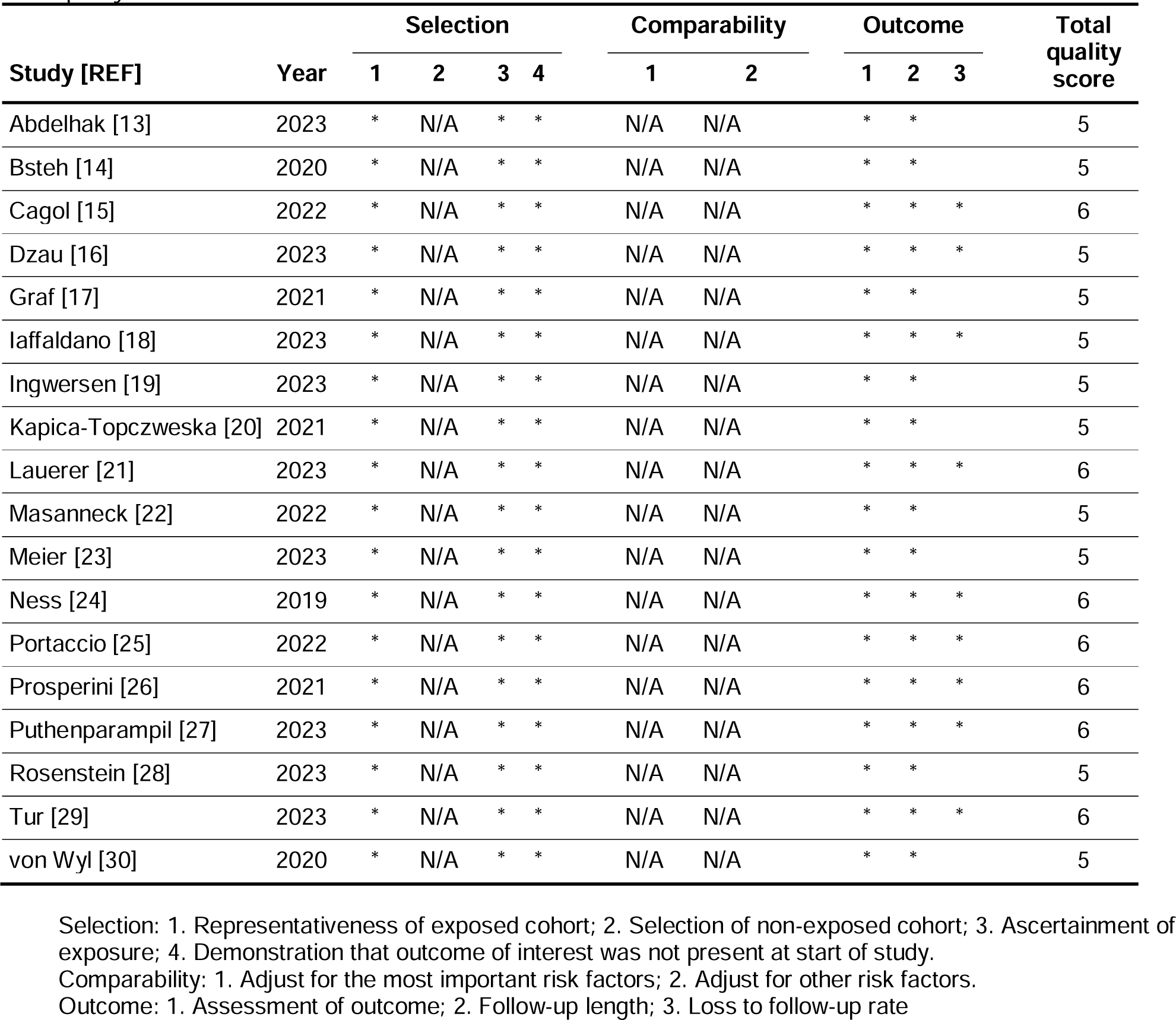
Assessment of methodological quality of included studies by the National Institutes of Health (NIH) quality assessment tool for observational cohort and cross-sectional studies.

### Definition of confirmed disability accrual

There was a relevant inconsistency in the definition of CDA across different studies, especially for PIRA. For the baseline score, a fixed (k = 11) [14]–[16],[18],[20],[21],[25]–[27],[29],[30] or roving (k = 9) [13],[17],[19],[22]–[24],[28] reference EDSS could be adopted. Regarding the event score, the stepwise clinically significant increase required for CDA was consistently ≥1.5 steps for a reference EDSS = 0 in all but one [17] of the included studies. An increase of ≥1.0 step was required for a reference EDSS of 1.0 to either 5.0 (k = 5) [21],[24],[26],[28],[29] or 5.5 (k = 14) [13]–[16],[18]–[20],[22],[23],[25],[27],[30]; accordingly, an increase of ≥0.5 step was required for a reference EDSS of either ≥5.5 or ≥6.0, respectively. Only one study adopted a different definition for CDA, requiring a ≥1.0 step-increase for a reference EDSS ≤3.0 and ≥0.5 step increase for a reference EDSS ≥3.5 [17]. According to different studies, the confirmation score was established at 3 (k = 2) [17],[19], 6 (k = 14) [13]–[16],[21]–[29] or 12 (k = 4) [13],[18],[20],[30] months after the event score. The sustained score was defined only in 7 studies at 12 months after the event score [14],[17],[24],[27] or at the end of observational follow-up [13],[16],[26]; the remaining studies did not report data on sustained score.

### Participants

The pooled cohort consisted in 52,667 patients followed for an observational period ranging from 2.4 to 12.1 years, corresponding to 415,825 patient-years; the **<u>TABLE 2</u>** shows their main characteristics. All studies comprised exclusively patients with RRMS, except for two ones including also small proportions with primary and secondary progressive MS [13],[23], i.e. 289 patients, corresponding to <1% of the pooled cohort.

**TABLE 2.** Summary of the main characteristics of included cohort studies (k = 20).

| Study Cohorts [REF] | Year | Sample size | Follow-up (years) | RAW event | PIRA event | Female:male ratio | Age (years) | Time since first symptom (years) | EDSS | DMTs |
| --- | --- | --- | --- | --- | --- | --- | --- | --- | --- | --- |
| Abdelhak-EPIC [13] | 2023 | 609 | 10.2 | 34 | 163 | 2.3 | 42.0 | 6.0 | 1.5 | Mixed |
| Abdelhak-SMSC [13] | 2023 | 1290 | 7.0 | 88 | 304 | 1.9 | 41.2 | 7.0 | 2.0 | Mixed |
| Bsteh [14] | 2020 | 171 | 4.0 | 28 | 40 | 2.7 | 35.2 | 6.1 | 1.5 | Mixed |
| Cagol [15] | 2022 | 516 | 3.2 | 35 | 60 | 2.1 | 41.4 | 9.5 | 2.0 | Mixed |
| Dzau [16] | 2023 | 10692 | 5.0 | 753 | 1199 | 2.4 | 32.2 | 0 | 2.0 | Mixed |
| Graf [17] | 2021 | 184 | 10.0 | 16 | 44 | 1.8 | 34.5 | 15.4 | 3.5 | High-efficacy |
| Iaffaldano [18] | 2023 | 16130 | 11.8 | 1720 | 7176 | 2.1 | 35.2 | 4.0 | 2.0 | Mixed |
| Ingwersen [19] | 2023 | 97 | 2.4 | 3 | 20 | 1.0 | 42.3 | 10.5 | 3.5 | High-efficacy |
| Kapica-Topczweska [20] | 2021 | 11632 | 5.0 | 475 | 569 | 2.4 | 36.0 | N/A | 2.0 | Mixed |
| Lauerer [21] | 2023 | 204 | 5.0 | 25 | 34 | 2.0 | 34.0 | 0.2 | 1.0 | Mixed |
| Masanneck [22] | 2022 | 46 | 5.6 | 11 | 19 | 3.6 | 32.8 | 0.5 | 1.5 | Mixed |
| Meier - Cohort 1 [23] | 2023 | 103 | 6.8 | 0 | 18 | 2.4 | 42.1 | 8.7 | 3.0 | Mixed |
| Meier - Cohort 2 [23] | 2023 | 252 | 3.1 | 4 | 39 | 1.6 | 44.3 | 9.9 | 3.0 | High-efficacy |
| Ness [24] | 2019 | 1959 | 2.0 | 150 | 166 | 2.7 | 41.6 | 7.3 | 2.0 | Mixed |
| Portaccio [25] | 2022 | 5169 | 11.5 | 922 | 1427 | 2.1 | 31.8 | 0.4 | 1.5 | Mixed |
| Prosperini [26] | 2021 | 687 | 12.1 | 175 | 76 | 2.3 | 30.8 | 4.6 | 1.5 | Mixed |
| Puthenparampil [27] | 2023 | 27 | 4.3 | 0 | 2 | 1.7 | 31.2 | 6.6 | 1.5 | High-efficacy |
| Rosenstein [28] | 2023 | 131 | 3.4 | 5 | 18 | 2.3 | 34.7 | 0.3 | 2.0 | Mixed |
| Tur [29] | 2023 | 1128 | 10.5 | 142 | 277 | 2.2 | 32.1 | 0 | 1.0 | Mixed |
| von Wyl [30] | 2020 | 1640 | 4.5 | 213 | 72 | 2.3 | 35.5 | 1.8 | 2.0 | Mixed |

A total of 4,799 patients experienced RAW, yielding a pooled event rate of 1.6 (95% CIs 1.1 to 2.1) per 100 patient-years; there was substantial heterogeneity across the included studies (Q_19_ = 613.7, *p* < 0.001; I^2^ = 99.4%), but without relevant publication bias (Kendall’s *τ* = 0.126, *p* = 0.460; Egger’s *Z* = –0.576, *p* = 0.564). A leave-one-out sensitivity analysis showed event rates for RAW ranging from 1.4 to 1.7 patient-years (*p* < 0.001), confirming that the pooled ES was not influenced by any single study.

A total of 11,723 patients experienced PIRA, yielding a pooled event rate of 3.1 (95% CIs 2.4 to 3.9) per 100 patient-years; there was substantial heterogeneity across the included studies (Q_19_ = 2618.8, *p* < 0.001; I^2^ = 99.5%), but without relevant publication bias (Kendall’s *τ* = 0.168, *p* = 0.319; Egger’s *Z* = 0.819, *p* = 0.412). A leave-one-out sensitivity analysis showed event rates for PIRA ranging from 2.9 to 3.2 per 100 patient-years (*p* < 0.001), confirming that the pooled ES was not driven by any single study.

We found that PIRA events accounted for 71.0% (95% CIs 70.3 to 71.7%) of all CDA events. Furthermore, 27.9% (95% CIs 26.4 to 29.5%) of relapses resulted in RAW events, as estimated on 11 articles [14],[15],[17],[19],[20],[22]–[24],[27],[28],[30] reporting data on the overall number of patients who relapsed during the observation period. There was no association between PIRA and RAW event rates (*p* = 0.210); however, RAW were almost completely suppressed, while PIRA accounted for about 82% of all CDA events, in cohorts including only patient under high-efficacy DMTs [17],[19],[23],[27] (**<u>FIG. 2</u>**).

**FIG. 2.**
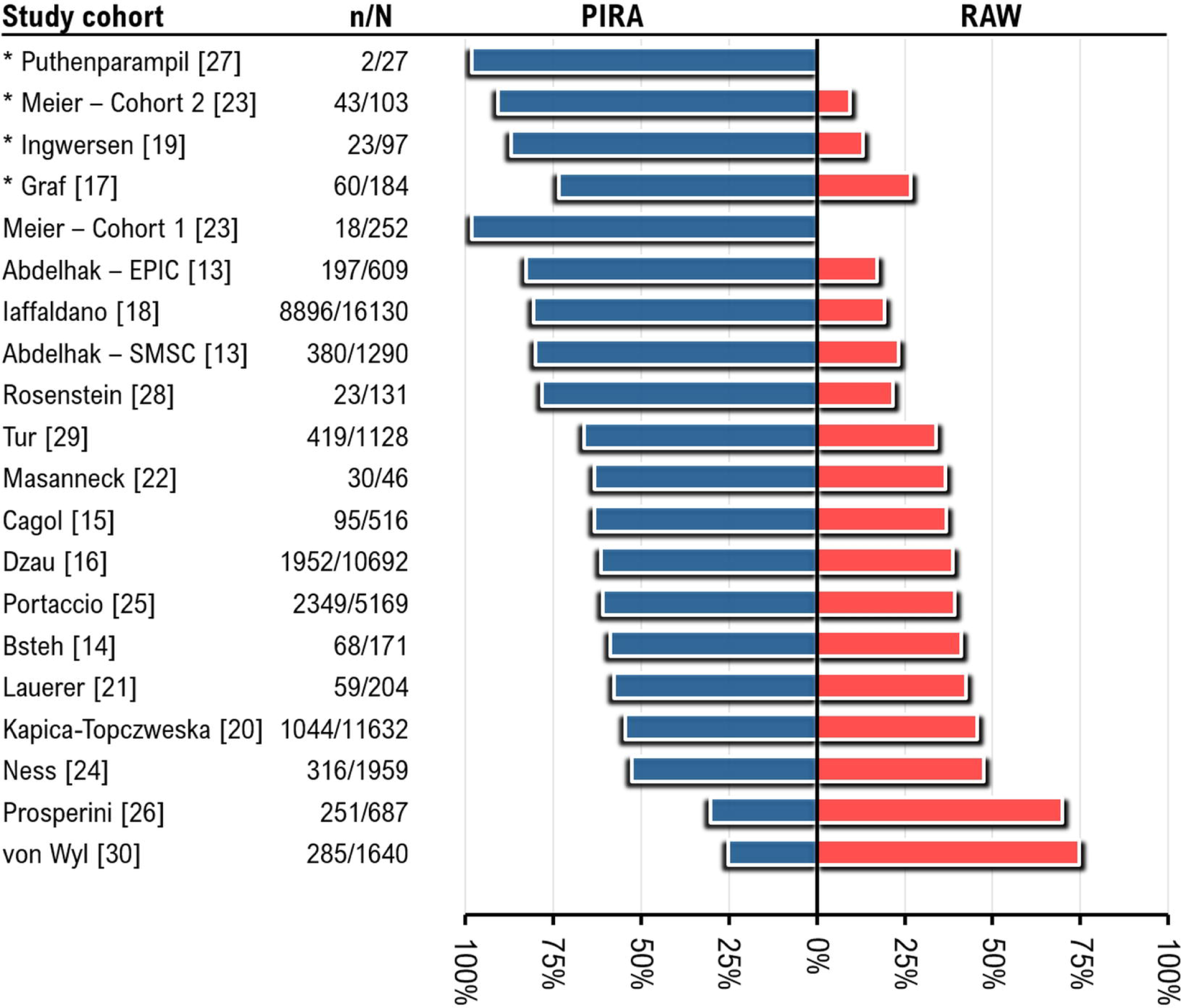
Proportions of patients experiencing different phenomena for confirmed disability accrual (CDA), i.e. progression independent of relapse activity (PIRA, blue bars) and relapse associated worsening (RAW, red bars) across different study cohorts (k = 20); each bar represents the 100% of patients experiencing CDA. *n/N: no. of patients experiencing CDA over sample size* *\* study cohorts including only patients under high-efficacy disease-modifying treatments (alemtuzumab, natalizumab, ocrelizumab, rituximab)*

### Determinants of PIRA and RAW events

Univariable meta-regression analyses showed no association between PIRA and RAW event rates, but there were different moderators for the two outcomes of interest (**<u>TABLE 3</u>**).

**TABLE 3.**
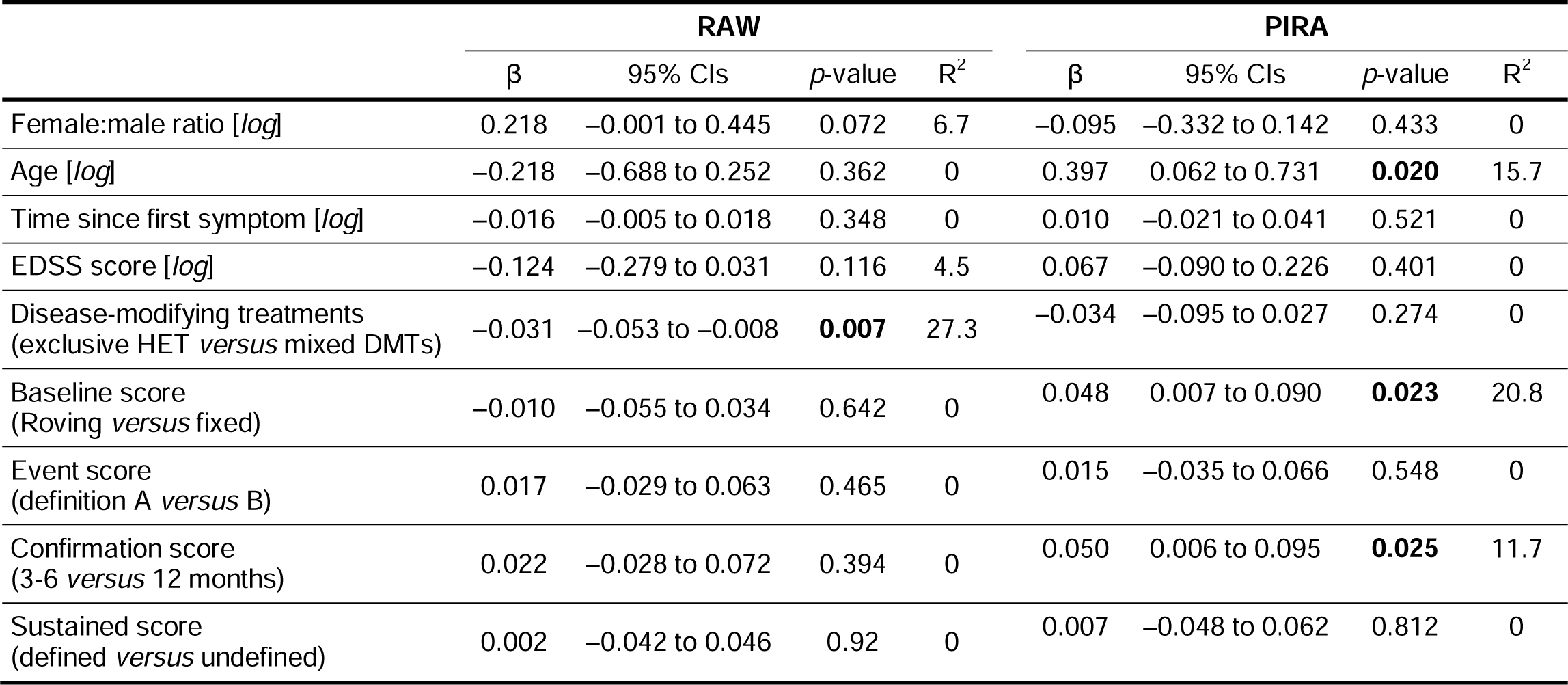
Meta-regressions exploring the effect of baseline clinical variables and criteria adopted to define confirmed disability accrual on RAW and PIRA event rates (double arcsine-transformed effect sizes).

|  | RAW |  |  |  | PIRA |  |  |  |
| --- | --- | --- | --- | --- | --- | --- | --- | --- |
| | $\beta$ | 95% CIs | *p*-value | R<sup>2</sup> | $\beta$ | 95% CIs | *p*-value | R<sup>2</sup> |
| Female:male ratio [*log*] | 0.218 | −0.001 to 0.445 | 0.072 | 6.7 | −0.095 | −0.332 to 0.142 | 0.433 | 0 |
| Age [*log*] | −0.218 | −0.688 to 0.252 | 0.362 | 0 | 0.397 | 0.062 to 0.731 | **0.020** | 15.7 |
| Time since first symptom [*log*] | −0.016 | −0.005 to 0.018 | 0.348 | 0 | 0.010 | −0.021 to 0.041 | 0.521 | 0 |
| EDSS score [*log*] | −0.124 | −0.279 to 0.031 | 0.116 | 4.5 | 0.067 | −0.090 to 0.226 | 0.401 | 0 |
| Disease-modifying treatments<br>(exclusive HET *versus* mixed DMTs) | −0.031 | −0.053 to −0.008 | **0.007** | 27.3 | −0.034 | −0.095 to 0.027 | 0.274 | 0 |
| Baseline score<br>(Roving *versus* fixed) | −0.010 | −0.055 to 0.034 | 0.642 | 0 | 0.048 | 0.007 to 0.090 | **0.023** | 20.8 |
| Event score<br>(definition A *versus* B) | 0.017 | −0.029 to 0.063 | 0.465 | 0 | 0.015 | −0.035 to 0.066 | 0.548 | 0 |
| Confirmation score<br>(3-6 *versus* 12 months) | 0.022 | −0.028 to 0.072 | 0.394 | 0 | 0.050 | 0.006 to 0.095 | **0.025** | 11.7 |
| Sustained score<br>(defined *versus* undefined) | 0.002 | −0.042 to 0.046 | 0.92 | 0 | 0.007 | −0.048 to 0.062 | 0.812 | 0 |

We found a more than 3.5-times higher RAW event rate in cohorts including patients under mixed DMTs (k = 16) than in those including patients under exclusive treatment with high-efficacy DMTs (k = 4), i.e. alemtuzumab, natalizumab, ocrelizumab, rituximab (β = 0.031, 95% CIs 0.008 to 0.053, *p* = 0.007) (**FIG. 3/A**).

**FIG. 3.**
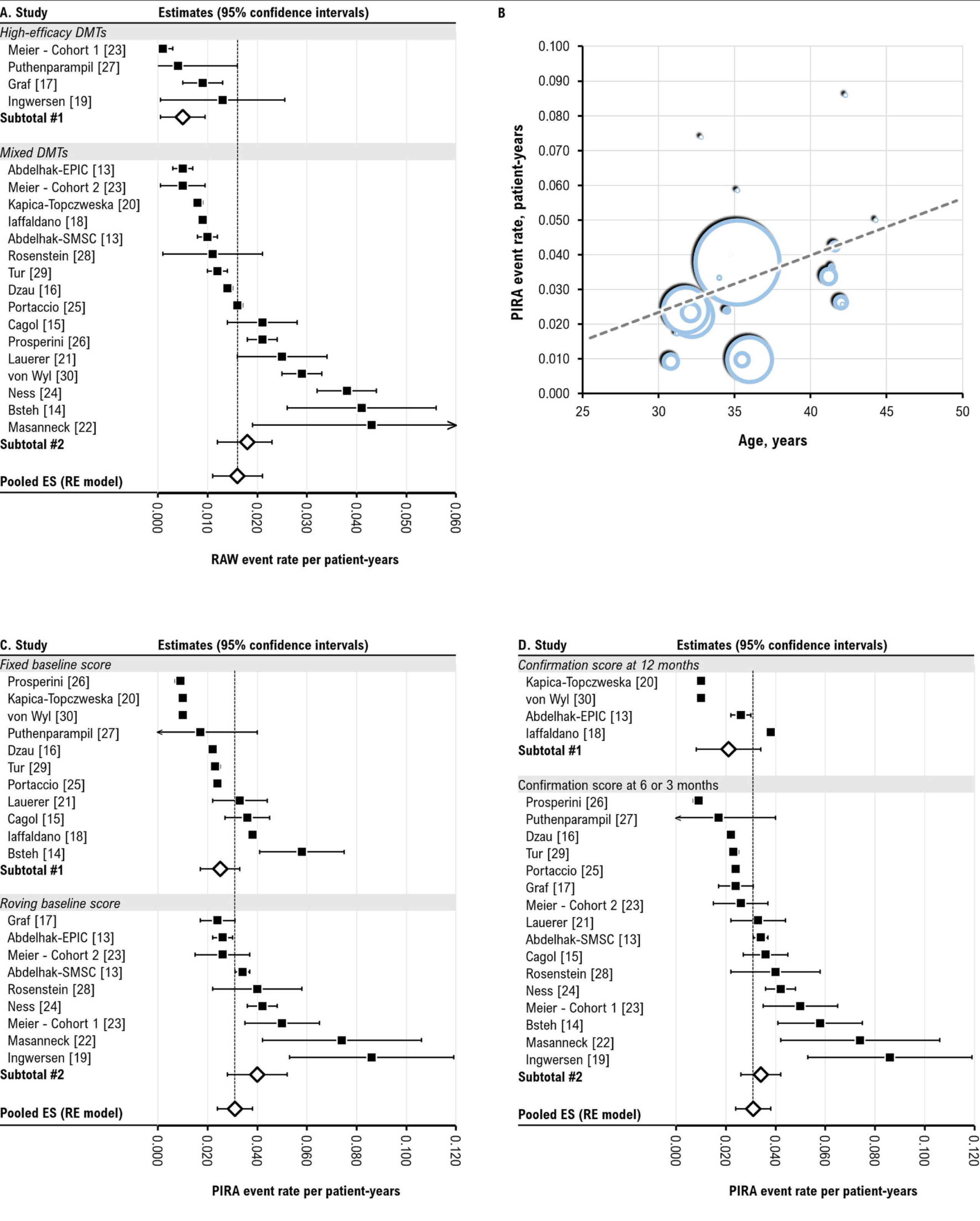
Forest plots for the subgroup analyses showing event rates of relapse associated worsening (RAW) by different types of disease-modifying treatments (A). Bubble plot for the association between event rates of progression independent of relapse activity (PIRA) and age at start of observation (B); each bubble represents a study cohort, with the bubble size proportional its sample size; the dotted lines indicates the interpolated regression line. Forest plots for the subgroup analyses showing event rates of progression independent of relapse activity (PIRA) by different criteria to define the baseline score (C) and the confirmation score (D).

The criteria adopted to define CDA did not influence the pooled RAW event rate. Older age at observation start was associated with higher rate of PIRA events (β = 0.397, 95% CI 0.062 to 0.731, *p* = 0.027); there was no further clinical factors influencing PIRA. We estimated a 1.6%-increase of PIRA event rate for each decade (**<u>FIG. 3/B</u>**), predicting ≥6 PIRA events per 100 patient-years at an age ≥54 years. The **<u>TABLE 4</u>** further illustrates how PIRA event rate estimates change at different ages. Additionally, we observed relevant differences in terms of PIRA event rates according to the criteria adopted to define CDA (**<u>FIG. 3/C and D</u>**). In more details, PIRA events were more frequent when adopting roving (k = 9) rather than fixed (k = 11) baseline score (β = 0.048, 95% CIs 0.007 to 0.090, *p* = 0.023), as well as confirmation score at 3 or 6 months (k = 16) rather than at 12 (k = 4) months (β = 0.050, 95% CIs 0.006 to 0.095, *p* = 0.025). Multivariable meta-regressions - by entering age at observation start and various combinations of the aforementioned significant covariates - confirmed the association between PIRA event rate and older age, regardless of the definition adopted for CDA in terms of baseline score and confirmation score (β = 0.395 and β = 0.478, *p* < 0.05).

**TABLE 4.** Estimates of progression independent of relapse activity (PIRA) event rates by different ages.

| Estimated PIRA event rate<br>*n* per 100 patient-years | Age (years) |
| --- | --- |
| 0.6 | 20 |
| 1.4 | 25 |
| 2.2 | 30 |
| 3.0 | 35 |
| 3.8 | 40 |
| 4.6 | 45 |
| 5.4 | 50 |
| 6.2 | 55 |
| 7.0 | 60 |
| 7.8 | 65 |
| 8.2 | 70 |

## DISCUSSION

### RAW and PIRA events in daily clinical setting

In this quantitative synthesis of 18 studies involving more than 50,000 patients followed for up to 12 years, we estimated less than two RAW events and approximately three PIRA events and per 100 patient-years in the real-world setting. Unlike post-hoc analyses of RCTs [6],[31] that assessed CDA events on the basis of composite score, real-world studies defined RAW and PIRA events solely based on EDSS change. This discrepancy may contribute to detecting more PIRA events in the experimental setting than in the real-world setting. Over 2-year follow-up, the proportions of patients with 3- and 6-month composite PIRA were 19.4% and 15.2%, respectively, in the pooled OPERA I and II population [6], and 13% experienced 3-month composite PIRA in the OPTIMUM study [36]. However, when considering only CDA assessed by EDSS change, our real-world PIRA estimate (3.1% *per annum*) becomes quite similar to those from the experimental setting, as the proportions of patients with 3- and 6-month PIRA were 5.7% and 4.5%, respectively, in the pooled OPERA I and II studies [6]. Consistently, the proportions of patients with 3- and 6-month PIRA were 5.4% and 4.2%, respectively, in the pooled ASCLEPIOS I and II population after 30 months of follow-up [31]. On the contrary, the difference in RAW were less relevant, being from 2.2% to 4.2% according to different RCTs [6],[31],[36] *versus* the 1.6% *per annum* estimated in this meta-analysis.

We also confirm that PIRA was the main reason for CDA even in real-world setting, and about one relapse out of three or four leads to RAW. Although there was no statistical association between RAW and PIRA event rates, we observed a concomitant reduction of RAW and increase in PIRA phenomena when neuroinflammation is strongly suppressed by high-efficacy DMTs (**<u>FIG. 3</u>**), as previously reported [1].

### Factors affecting RAW and PIRA events

The event rates for RAW and PIRA were affected by different moderators, substantiating the notion of different mechanisms underlying these two phenomena. In particular, RAW has been observed more often in patients with signs of MRI activity, such as contrast-enhancing and new T2-hyperintense lesions [1],[26]. On the other hand, PIRA has been relates with brain atrophy [15], spinal cord damage (in terms of lesions and atrophy) [21],[26],[37], retinal thinning [14], biomarkers of neuroaxonal and astrocyte damage, such as serum neurofilament light chain and glial fibrillary acidic protein, respectively [13],[23].

According to our findings, age is the only determinant of PIRA, with a 1.6%-increase of event rate for each decade. The effect of age on PIRA event rate remained unaltered even after adjusting for different criteria adopted to define CDA. Notably, the PIRA event rate was largely influenced by such criteria. A recent systematic review already raised the concern about the inconsistency of CDA definition as a relevant source of variability requiring harmonization [7]. Using meta-regression equations, we were able to quantify this variability in 1.5 and 1.3 more events *per annum* when a roving rather than fixed EDSS was adopted as baseline score and when the confirmation score was established at 3 or 6 rather than 12 months, respectively.

We also found that treatment with high-efficacy DMTs strongly suppresses RAW, while there was no effect on PIRA by DMTs [7]. This latter finding somewhat conflicts with post-hoc analysis of two RCTs where ocrelizumab and ofatumumab reduced the risk of PIRA *versus* active comparators interferon beta and teriflunomide [6],[31]. However, the effect of high-efficacy DMTs on composite PIRA was considerably smaller than that on RAW, even losing its statistical significance on PIRA assessed as 24-month confirmed EDSS change in the pooled OPERA I and II population [6]. In a post-hoc analysis of the OPTIMUM study, there was no difference between ponesimod and teriflunomide on composite PIRA [36].

Notably, while the findings from afore mentioned post-hoc studies were based on a direct comparison, our study design only allows indirect comparison that is biased by cross-RCT differences in baseline patients’ characteristics and in outcome definition.

### Clinical and methodological implications

We emphasize the need for a cautious approach in handling and interpreting PIRA outcomes, especially in the real-world setting. Our findings highlight that the PIRA event rates are largely contingent on the criteria employed to define CDA. Thus, we advocate for the appropriate harmonization of CDA definitions to ensure consistency and precision in evaluating PIRA outcomes [7]. Another issue concerns the low inter-rater reliability of EDSS, especially at the lower scores (calculated only on individual functional systems), that is only partially mitigated by subsequent confirmation visits. Older individuals with and without MS demonstrated significant disability on the EDSS, indicating that factors beyond MS may affect the score, as demonstrated in a small exploratory study involving individuals over 55 years [38]. This suggests that certain functional systems assessed by EDSS, like balance, sensation, and sphincter function, may be partially influenced by factors other than MS, especially those associated with age-related deterioration [38]. As the primary influence on PIRA appears to be age, prompting concerns regarding whether specific CDA events are more driven by disease-related or age-related mechanisms. Composite scores including not only EDSS, but also assessments for manual dexterity, walking speed and cognitive functions could enhance the precision in PIRA detection. However, their adoption in clinical settings seems impractical. Additionally, it is important acknowledge that stopwatch tests, like 25-FWT, 9-HPT, SDMT, are subjected to learning effect [39].

A crucial aspect to address is the statistical approach to RAW and PIRA. If we assume that these two phenomena can co-occur in the same individual, a certain mutual exclusivity way exist, as the occurrence of one event might preclude the other one. Specifically, only CDA event within 30 days before and 90 days after the onset of a relapse should be considered for classification to PIRA [7]. Hence, a competing risk analysis, which estimates the marginal probability of an event in the presence of competing events, appears to be more appropriate than the standard time-to-event analysis [40].

Moreover, all studies employed time-to-event analysis, interpreting their main results as number of patients with at least one RAW or PIRA event. However, statistical models based on count of events (e.g. Poisson or negative binomial regression) could be more suitable, given that the same individual can experience two or more RAW/PIRA events throughout their lifetime. Nevertheless, count-based models require a number of assumptions which may be theoretically violated in the context of RAW and PIRA, primarily concerning the independence of events and the absence of zero inflation, i.e. the likelihood of observing zero events must be comparable to the likelihood of observing one or more events.

### Limitations

Our meta-analysis faces limitations associated with synthesizing observational data from pooling cohorts, introducing within-study and across-study biases that contribute to relevant statistical heterogeneity and inconsistency.

There is a relative risk of partial data duplication in three article from Switzerland [13],[15],[23] and two articles from Italy [18],[25]. Despite this concern, opted to retain all available data due to the substantial difference in sample sizes. Moreover, this meta-analysis failed to estimate the rate of true PIRA, defined as CDA events in absence of both relapses and magnetic resonance activity, a metric rarely reported in the selected studies. Another drawback is the inability to adjust the estimates for the number of assessments, as the frequency of visits can significantly influence the detection of outcomes. While our statistical methodology is robust, our findings may not be directly applicable to individuals due to the risk of ecological fallacy, i.e. the associations identified in pooled data does not necessarily reflect what happens at the patient level. Validation of ecological hypotheses at the individual level would require individual-level analysis, which is unfeasible due to the unavailability of raw datasets.

### Conclusions

We reaffirm that PIRA predominantly contributes to CDA events detected in real-world setting, even in the early stages of MS, whereas RAW emerges as a less frequent phenomenon, likely thanks to the widespread use of effective DMTs. However, it is crucial to acknowledge the inherent challenges in the detection and statistical interpretation of PIRA outcomes, raising concerns about potential biases despite efforts towards harmonization [7]. The observation of both RAW and PIRA events even in the earlier disease stage, even though the intrinsic and perhaps overwhelming afore mentioned limitations, underscores the concept that MS may be viewed as a *continuum*. This challenges the strict definition of disease phenotypes and aligns with the recent understanding of a prolonged preclinical phase, emphasizing the intricate interplay between lesion localization, frequency and severity of focal lesions, magnitude and progression of diffuse damage, age-related processes, and the structural and functional reserve available.

## Supporting information

PRISMA checklist

## Data Availability

An Excel spreadsheet containing all data of included studies is available as an Appendix.

## ACKNOWLEDGEMENTS

We thank Dr. Philip Albrecht, Dr. Pietro Iaffaldano, Dr. Katarzyna Kapica-Topczewska, Dr. Lars Masanneck and Dr. Marco Puterampil for data sharing.

## AUTHOR CONTRIBUTIONS

Conception and design of the study, drafting a significant portion of the manuscript/figures: LP, SR, SH. Acquisition and analysis of data, revision of manuscript content: LP, SR, CT.

Supervision and drafting the final version of the manuscript: LP, SH, CG.

## DATA SHARING STATEMENT

An Excel spreadsheet containing all data of included studies is available as an Appendix.

## ETHICAL STATEMENT

not applicable.

